# The hyperacute plasma proteome reports injury severity and defines distinct trauma endotypes

**DOI:** 10.64898/2026.01.31.26345281

**Authors:** Sara Masarone, Gerard Hernández Mir, Jennifer Ross, Jason Pott, Karim Brohi, Michael R. Barnes, Daniel J. Pennington

**Affiliations:** Centre for Immunobiology, Blizard Institute, Queen Mary University of London; London, E1 2AT, UK; Alan Turing Institute; London, NW1 2DB, UK; Centre for Trauma Sciences, Blizard Institute, Queen Mary University of London; London, E1 2AT, UK; Centre for Translational Bioinformatics, William Harvey Research Institute, Queen Mary University of London; London, EC1M 6BQ, UK

## Abstract

The host response to traumatic injury is varied and unpredictable. Patients with subjectively similar injuries progress along divergent clinical paths, from uncomplicated recovery to extended hospitalisation, multiple organ dysfunction, and life-long ill health. The temporal certainty of trauma nonetheless provides immediate opportunity to predict clinical trajectory and intervene therapeutically. We demonstrate using machine-learning that the hyperacute plasma proteome can function as a physiology-driven indicator of injury severity. Moreover, it identifies two serious-injury endotypes that differentially predict unfavourable clinical trajectories. Notably, a prominent neuronal guidance protein signature, that reports vascular dysfunction and immune activation, together with an anti-coagulation/pro-fibrinolytic state, identify those patients who progress to adverse clinical outcomes. Together, these mechanistic insights into immediate host responses to serious trauma reveal tractable targets for future therapeutic interventions.

---

Trauma is a leading cause of morbidity and mortality worldwide (*1*). Clinical trajectories are set within minutes, determined by injury severity and seemingly, by differential host responses to tissue damage (*2*); individuals with similar anatomical descriptions of injury progressing on divergent clinical paths, ranging from complication-free recovery to prolonged multiple organ failure, recurrent infection, extended intensive care requirements, and death (*3*). To better understand this, we considered two competing hypotheses; that the likelihood of having an adverse clinical outcome, after severe injury, either correlates directly with increasing injury severity; or is dependent on distinct host responses emanating from similar injury severity states. Clinically, the gold standard for assessing injury severity is the injury severity score (ISS) that is derived using the abbreviated injury scale (AIS) (*4–6*). This score is assigned retrospectively and is based on findings from diagnostic radiology or direct surgical visualisation. Although ISS somewhat relies on subjective clinical assessments, it does predict adverse clinical outcomes with reasonable accuracy, albeit not in a timeframe that is operationally useful. Thus, we reasoned that a biologically-driven objective indicator of injury severity, derived from the hyperacute plasma proteome of seriously-injured individuals, may better predict immediate clinical trajectories. Moreover, we postulated that these “serious-injury” signatures could also reveal the distinct physiological perturbations that describe traumatic injury, that correlate with poor prognosis in the clinic, and that represent potential future targets for therapeutic intervention.

## The hyperacute plasma proteome reports injury severity

Blood plasma was obtained in the “hyperacute window” (<2hr from injury), from 414 patients with blunt trauma but clinical absence of major traumatic brain injury, recruited through the ACIT study (*7*). Patients were selected on graded ISS; low (ISS 0-3; n=73), mild (ISS 4-8; n=77), moderate (ISS 9-15; n=79), severe (ISS 16-24; n=77) or critical (ISS >24; n=108). As expected, the probability of adverse clinical outcomes rose significantly with ISS, and correlated at admission with higher white cell count, and increased base deficit and lactate levels (**table S1**).

A proteomic dataset from double-spun plasma was then generated using the aptamer-based Somalogic SomaScan© technology for expression of 4979 protein analytes (*8*). Exploratory data analysis indicated a general increase in protein levels in critical patients compared to those with low ISS (4102 & 192 proteins, significantly elevated or decreased, respectively). Many of these proteins were also significantly correlated with increasing ISS across the patient cohort (**fig. S1**), including those already mechanistically implicated in adverse consequences of severe trauma, such as PROC (activated protein C) (*9*) and IL-6 (*10*) (**fig. S1**).

We next used supervised machine-learning (ML) to identify key proteomic signatures that predict injury severity. We utilised XGBoost (eXtreme Gradient Boosting - a scalable gradient-boosted decision tree ML library), to generate proteomic models that predict critical injury, where critical injury was defined as being above a particular ISS threshold; from ISS>24 (that is used clinically to define “critical” injury) to ISS>33. Assessment of AUC values for receiver operator curves demonstrated high model performance (0.85-0.90) (**Fig. 1A**). In addition, the absence of a pronounced improvement in model performance as we incrementally increased ISS threshold suggested that a sudden step-switch from a non-critical to critical proteomic signature was unlikely to exist.

**Figure 1.**
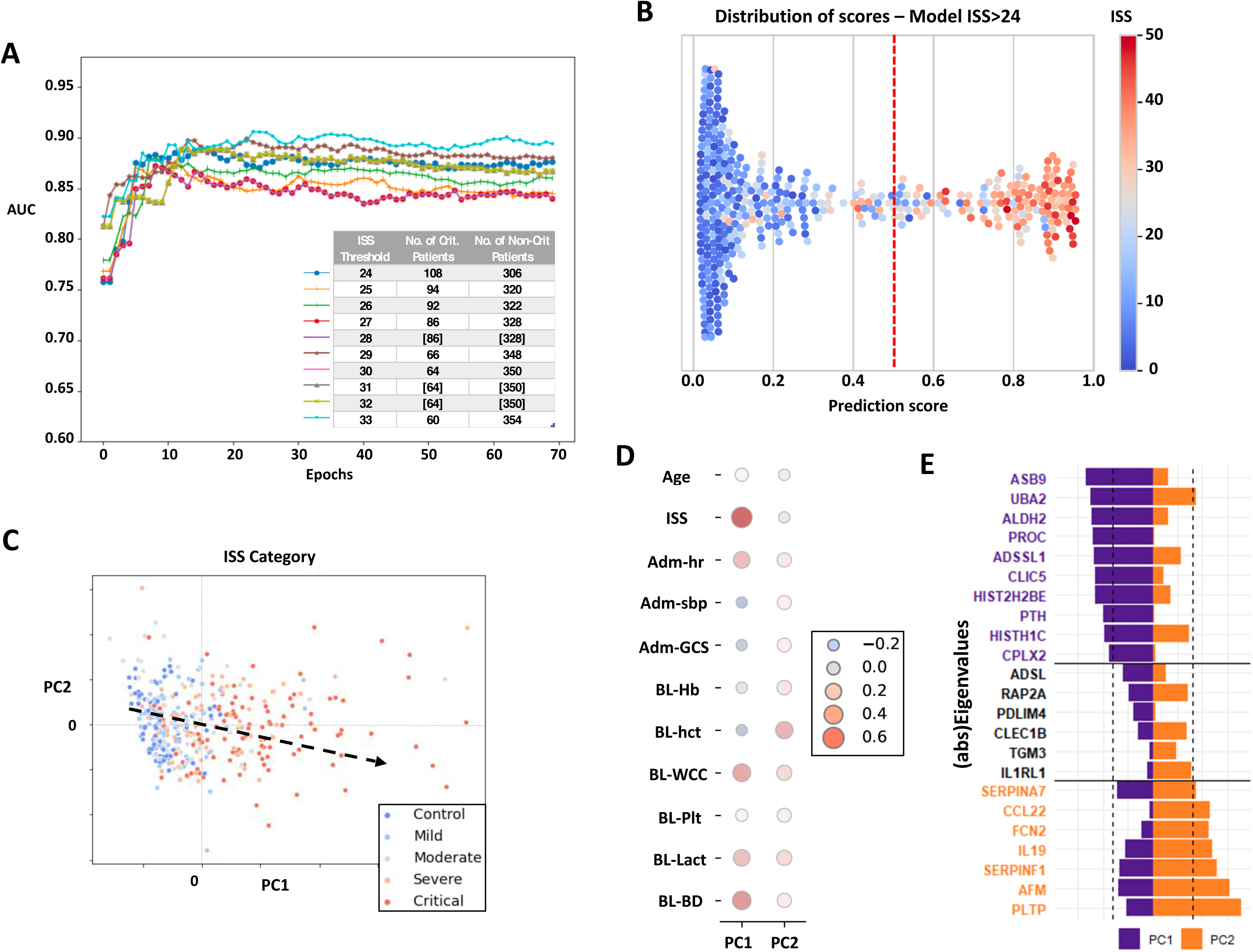
The hyperacute plasma proteome reports injury severity. **(A)** Performance (Receiver’s Operator Curve) of models (70 epochs shown) that predict critical injury at different ISS thresholds (from ISS>24 to ISS>33). The table shows the number of patients that belong to each group (critical vs “non-critical”) for each threshold. **(B)** Distribution of prediction scores from model built using only the 23 ML-derived proteins and ISS>24 as threshold. Actual ISS is indicated by colour. Dotted line defines boundary between critical and “non-critical” with critical patients having prediction scores >0.5. **(C)** PCA based on expression of the 23 M-L-derived proteins in our patient cohort. Arrow shows direction along which severity segregates. **(D)** Spearman correlations between PC1 and PC2 (from (A)), and clinical variables. Stronger correlations are indicated by larger dots. Positive correlations are red, negative correlations are blue. **(E)** Table showing values for each protein for each Principal Component (from (C)). Purple and orange text signify PC1-dominant and PC2-dominant proteins, respectively.

An explainable ML approach allows for feature selection; here, proteins that make significant discriminative contributions to the predictive capacity of each of the ten models trained on critical injury thresholds ISS>24 to ISS>33. This identified 23 proteins; ADSL, ADSSL1, AFM, ALDH2, ASB9, CCL22, CLEC1B, CLIC5, CPLX2, FCN2, HIST1H1C, HIST2H2BE, IL-19, IL1RL1, PDLIM4, PLTP, PROC, PTH, RAP2A, SERPINA7, SERPINF1, TGM3, & UBA2 (**table S2**). We then used only these 23 proteins to construct a final ML model whose function predicted critical injury at the ISS>24 threshold (ML_(23)_^ISS>24^ model). Precision (the proportion of correctly predicted positive instances among all instances that were predicted as positive), and recall (the proportion of correctly predicted positive instances among all instances that are truly positive), for the ML_(23)_^ISS>24^ model were high, at 0.82 and 0.80, respectively, which resulted in an F1 score of 0.81 and AUC of 0.88 (**table S3**). Moreover, the model’s predictive capacity correlated well with ISS (**Fig. 1B**), demonstrating that such an approach could be used to generate an objective proteomic-based “plasma injury score”. Indeed, in a test-set of 125 patients, admission characteristics and probabilities of adverse clinical outcomes were very similar between patients with actual ISS values above 24, and those that were predicted as having an ISS>24 using the ML_(23)_^ISS>24^ model (**table S4**).

Overlaying ISS categories on a PCA plot of the 414 patients driven by expression of the 23 ML-selected proteins demonstrated that patients with increasing ISS could be segregated effectively (**Fig. 1C**). This was confirmed with a correlogram of PC1 and PC2 against clinical admission data that revealed ISS as a dominant component of PC1 (**Fig. 1D**). Notably, Eigenvectors for this PCA plot demonstrated that PC1-dominant proteins (e.g., ADSSL1, ASB9, HIST2H2BE, and UBA2), were those that featured prominently in the list of top 50 ISS-correlated proteins (**Fig. 1E and fig. S1 and S2**). However, PC2-dominant proteins (e.g., IL-19, PLTP, SERPINA7, and SERPINF1), that are linked to both hemostasis and immune function, did not feature in this list, or in the list of top differentially expressed proteins between critical and control patients (ISS>24 *vs* ISS 0-3), suggesting that potentially important, but less obvious, additional components of injury severity may have been revealed by our ML approach.

## Damage-associated proteins are differentially elevated in critically injured patients

A ML-model that predicts critical injury is likely to identify index proteins from key physiological processes that are perturbed after severe trauma. To characterize these indexed processes, we clustered the 4979 SomaScan©-associated proteins on a tSNE plot, based on their correlated expression across our 414 patients. This identified 20 clusters (**Fig. 2A**), that we linked to potential physiological functions using KEGG pathway analysis (**Fig. 2B, and table S5**). Notably, the 23 ML-selected proteins located particularly in clusters 2, 9, and 17, that featured PC1-dominant proteins, and clusters 8, 10, and 19 that featured PC2-dominant proteins (**Fig. 2B**). Clusters 2, 9, and 17 also contained the majority of the top 200 differentially-expressed proteins between critical and low-ISS patient cohorts (**Fig. 2C**). Moreover, when average protein levels for patients from specific ISS categories were displayed relative to the average protein level of that protein in all patients, proteins in clusters 2, 9 and 17 were increased to a greater extent than proteins in other clusters in critically-injured patients, while proteins in clusters 8, 10 and 19 decreased (**Fig. 2D**). These notable changes, that correlated with increasing ISS, were also evident on a heat-map of ML-selected proteins in which proteins from the same cluster were displayed together (**Fig. 2E**). Collectively, these data suggest that increasing injury severity (as benchmarked against ISS), is characterised by an intracellular proteome (e.g., muscle-specific enzyme ADSSL1) that reports tissue damage. However, these analyses also suggest more subtle physiological changes, related to hemostasis (cluster 19), and immune function (clusters 8 & 10), that may modulate clinical trajectory and probability of adverse clinical outcomes.

**Figure 2.**
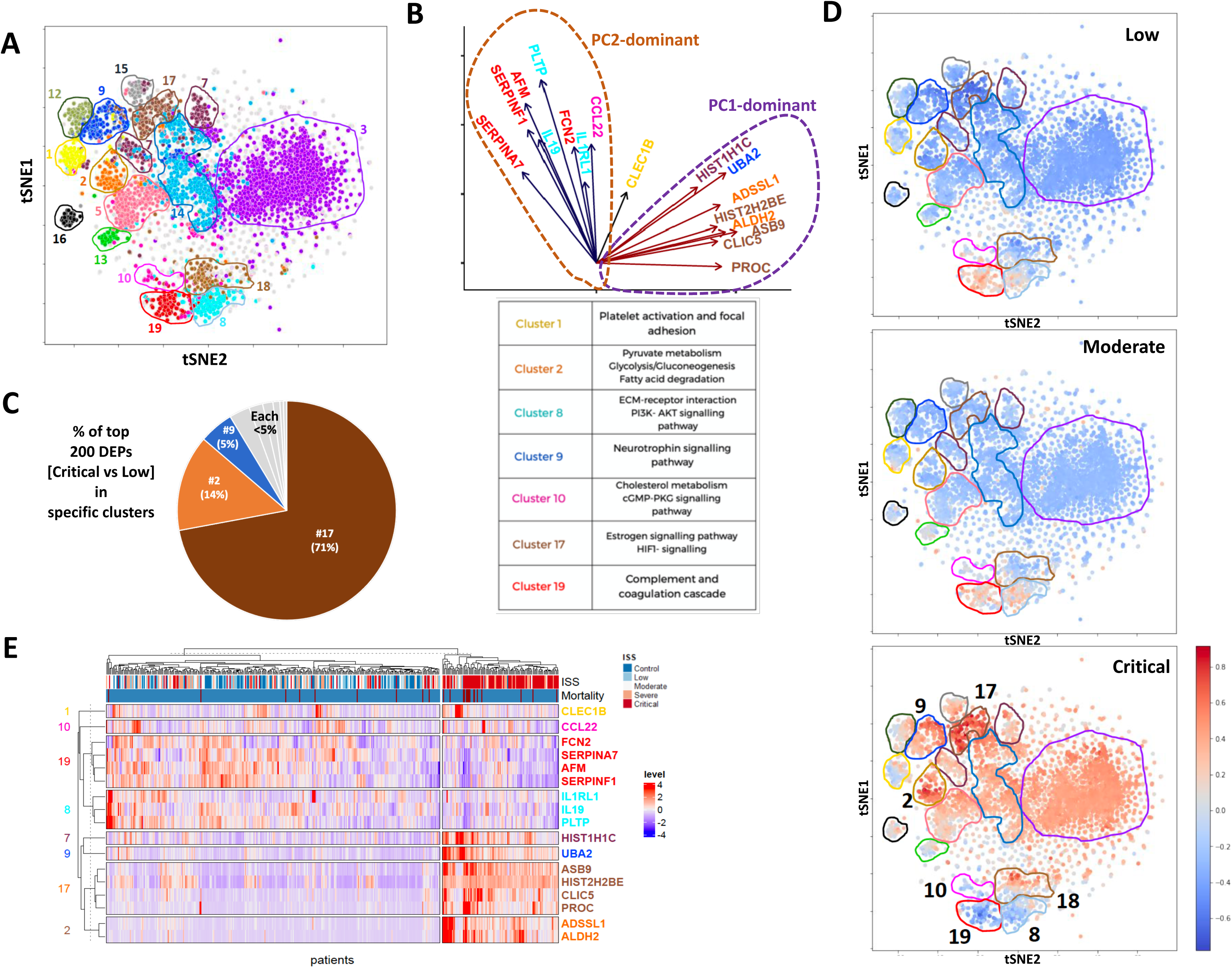
Damage-associated proteins are differentially elevated in critically injured patients. **(A)** Low dimensional visualisation (t-SNE plot) showing proteins clustered (K-means) based on expression in our 414-patient cohort. **(B)** Major eigenvectors (from PCA in figure 2) with proteins coloured by their corresponding cluster in (A). The table summarises the main pathways associated with these proteins. **(C)** Pie chart displaying the percentage of top-200 differentially expressed proteins (ISS>24 *vs.* ISS=0-3) belonging to each K-Means cluster identified in (A). **(D)** t-SNE plot representations for ISS-defined low, moderate and critical patients, showing average expression of all proteins for each group relative to the mean in all patients. **(E)** Heatmap showing expression of top 17 proteins in each trauma patient, highlighting the relationship between ISS scores and protein expression.

## CPLX2 identifies two serious-injury endotypes

We next considered the concept of serious-injury *endotypes*; defined as distinct injury-associated phenotypes explained by underlying pathobiological mechanisms. To explore this, we analysed Shapley values (a model’s explainability values), that quantify the importance of each protein to the predictive capacity of the ML_(23)_^ISS>24^ model for each patient. A SHAP plot revealed that, across the cohort, high expression of ADSSL1, CPLX2, CLIC5, ASB9, PTH and HIST2H2BE, but low levels of CCL22, HIST1H1C and SERPINA7, had the most positive impact on predicting ISS>24 for the ML_(23)_^ISS>24^ model (**Fig. 3A**). However, force plot analysis, that shows the combination of major protein contributions that push the model’s prediction score above 0.5 for that individual (i.e., that predicts critical injury for that person), can be different for different patients; for example, being driven by aberrant expression of SERPINF1, CLIC5 and CCL22 in one person, but by ADSSL1, HIST1H1C and CPLX2 in another (**Fig. 3B**).

**Figure 3.**
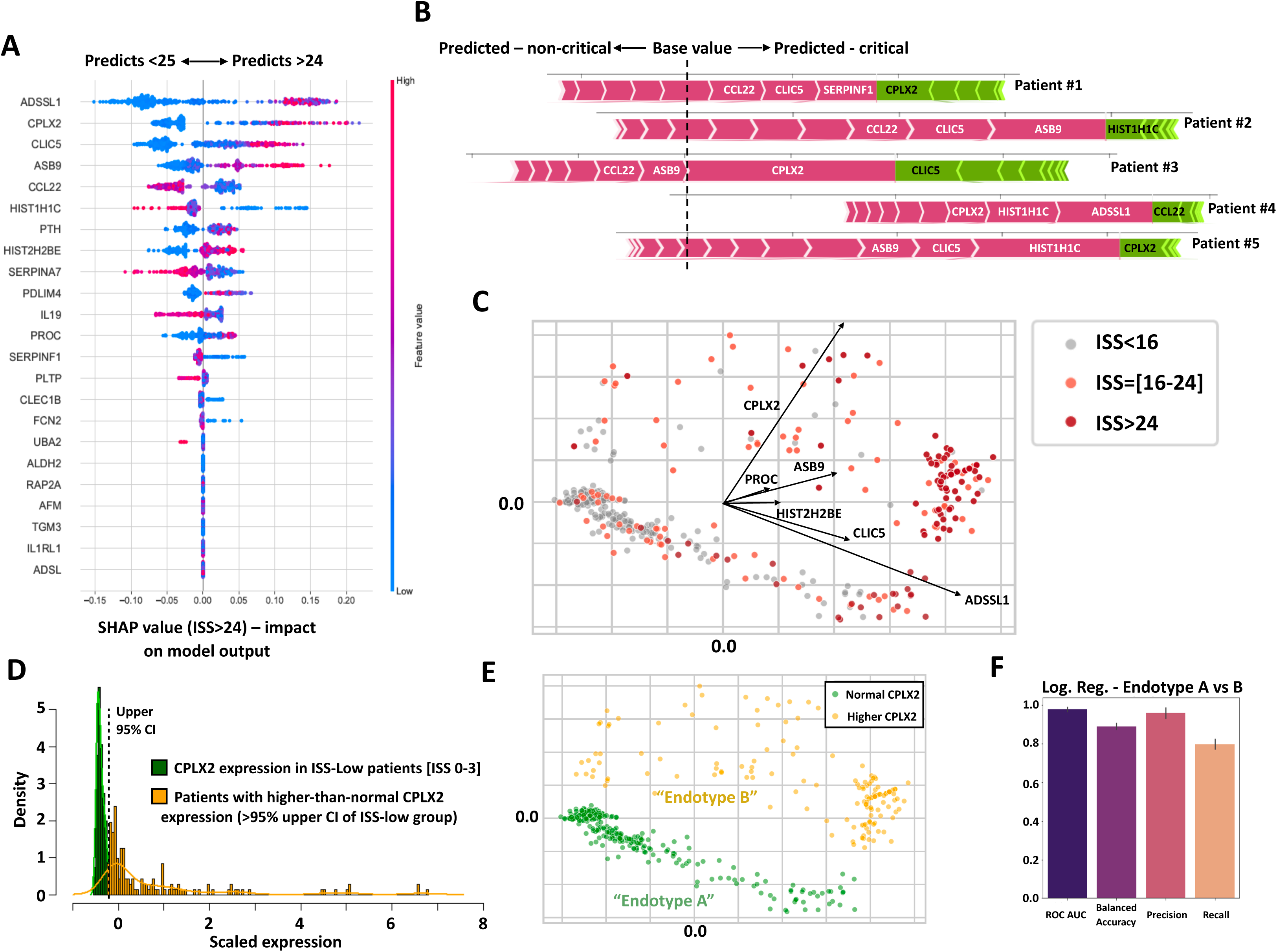
CPLX2 identifies two serious-injury endotypes. **(A)** Shap summary plot showing a global overview of model using 23 ML-derived proteins and critical injury threshold ISS>24. Plot indicates impact and contribution (positive or negative) of expression level of each protein to the final prediction score. Colour of dots represents relative plasma protein levels. **(B)** Force plots showing model predictions for individual patients. Plots show contributions to prediction score of most dominant proteins in that individual. **(C)** PCA of the Shap values generated by the model utilizing the 23 ML-derived proteins to predict critical injury (i.e., ISS>24). Patients are represented by dots and are coloured to represent ISS. **(D)** Histogram and density plot of CPLX2 expression. Dark green denotes expression of CPLX2 in low-ISS patients (with ISS=0-3). Yellow denotes patient CPLX2 expression outside this range. Green/yellow boundary is determined using the 95^th^ percentile of expression of CPLX2 low-ISS patients. **(E)**. PCA (from (C)) coloured by CPLX2 expression. Patients with CPLX2 within the “normal” range (as defined in (D)) are coloured green (denoted “endotype A”), patients outside that range are coloured yellow (denoted “endotype B”). **(F)** Result of logistic regression analysis when predicting the two endotypes (A *vs*. B).

We next reasoned that endotypes might be revealed on a PCA plot that would cluster patients with similar Shapley value contributions to their predictive score for injury severity. Conversely, if every prediction of critical injury involved a different combination of Shapley value contributions, no clusters would be apparent. Unsurprisingly, most of the patients with lower injury severity scores (i.e., ISS<16) clustered together (**Fig. 3C**). Increasing ISS correlated with PC1, with most of the critical patients having high PC1 values. However, there appeared to be two groupings of more-injured patients (ISS>15); one towards the bottom of the plot, co-localising with most of the less-injured patients, and a second towards the top of the plot. To interrogate these groupings further, eigenvectors for the PCA were calculated. This suggested that Shapley values relating to 6 of the 23 proteins were particularly dominant. Five of these, ADSSL1, CLIC5, ASB9, PROC and HIST2H2BE, largely contributed to PC1 and increasing ISS (**Fig. 3C, and fig. S3**), consistent with our earlier analyses. By contrast, CPLX2 was the sole major driver of PC2.

Complexin-2 functions in synaptic vesicle exocytosis and is normally found at very low levels in the plasma of healthy individuals. Notably however, 143 patients (across a range of ISS), had significantly raised CPLX2 levels, that surpassed the 95^th^ percentile boundary for CPLX2 levels in our low-ISS [ISS 0-3] group (**Fig. 3D, and fig. S4**). When mapped onto the PCA of Shapley values generated by the ML_(23)_^ISS>24^ model, these 143 individuals clearly segregated as an upper group, that we term “endotype-B”, distinct from a lower cluster of patients, termed “endotype-A” (**Fig. 3E**). A logistic regression analysis (with 5-fold cross-validation), trained to predict whether a patient was endotype-A or endotype-B, showed good performance with high precision and recall and an ROC AUC of >0.95 (**Fig. 3F**). Thus, these data reveal (at least) two serious-injury endotypes; the first (endotype-A) with plasma-associated proteomic features consistent with patients with lower ISS, and a second (endotype-B) in which protein levels are significantly perturbed.

## Endotype-B patients display a pronounced neuronal guidance proteomic signature

To explore the two serious-injury endotypes further we next compared patients from each endotype with an ISS>15 (i.e., severe, and critical patients only). A comparison of average age, ISS, systolic blood pressure, white cell count, lactate level and base deficit at admission did not reveal significant differences (**table S6**). By contrast, endotype-B individuals had much higher plasma protein levels than endotype-A patients (**Fig. 4A**). The top 200 of these up-regulated proteins mapped largely to clusters 2, 9, and 17 that we had previously identified as indicative of tissue damage (**fig. S5**). However, more than 35 proteins in cluster 18 were also significantly elevated (**Fig. 4B, and fig. S5**). Notably, KEGG and Reactome pathway analyses of these proteins identified a prominent “axon guidance” signature (**fig. S6**). The axon, or neuronal, guidance protein (NGP) family has multiple functions aside from patterning the embryonic neural network, and is characterised by five major groups: the netrins, semaphorins, ephrins, repulsive guidance molecules (RGMs), and slits (*11–13*). Ligands or receptors from the first four of these featured prominently in the endotype-B signature: netrin-1 receptor, UNC5B; semaphorin SEMA7A; ephrin receptor, EPHB6; and RGMB (**Fig. 4C**). Indeed, markedly elevated levels of proteins from the NGP network was a defining feature of endotype-B individuals (**fig. S7**), that did not appear to simply correlate with indicators of neuronal damage, such as AIS-Head scores that were not notably different between endotype-A and endotype-B patients with severe or critical injuries (i.e., ISS>15) (**fig. S8**).

**Figure 4.**
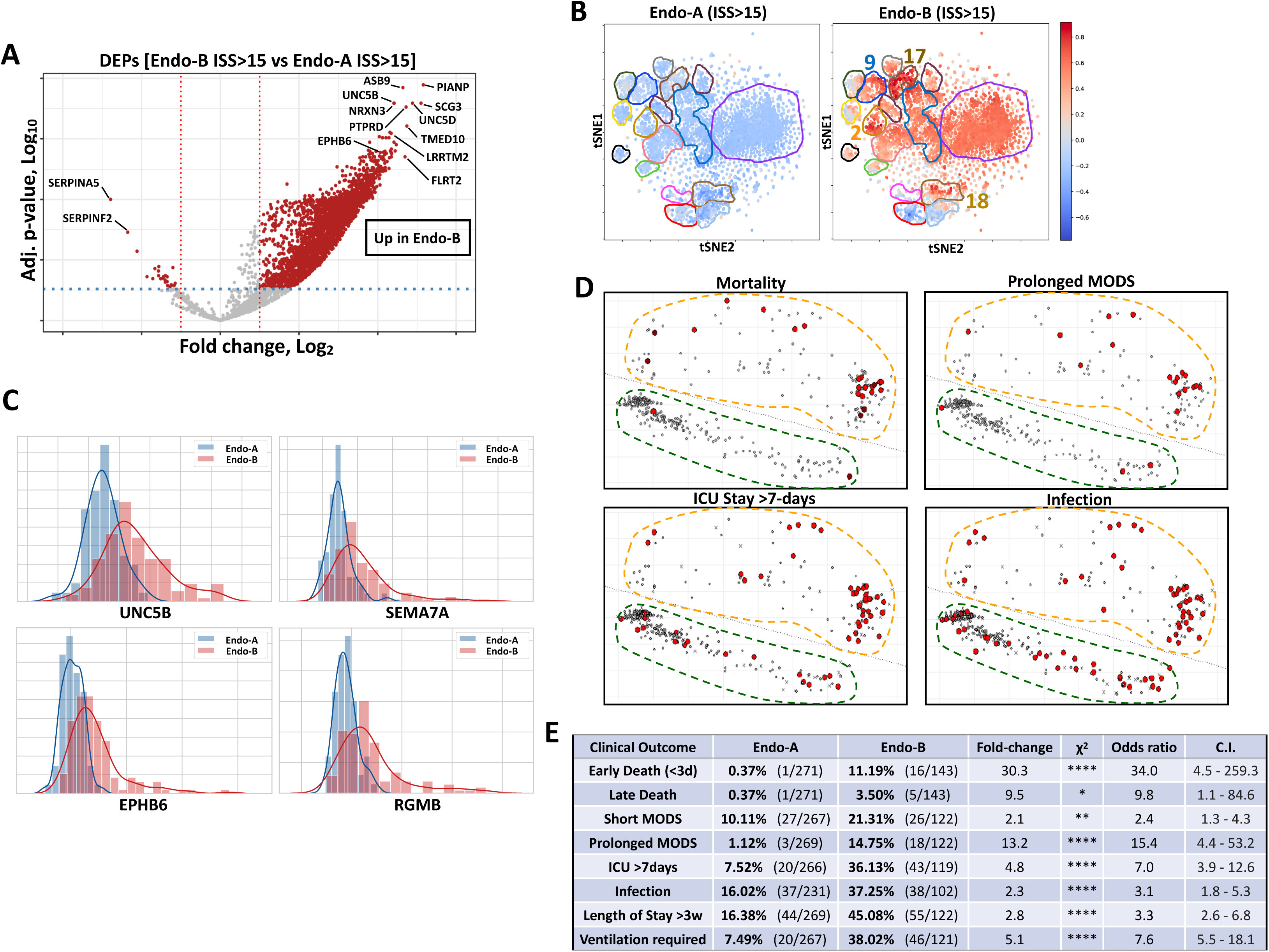
Endotype-B patients display a pronounced NGP signature and have greater likelihood of adverse clinical outcomes. **(A)** Volcano plot of differentially expressed proteins between endotype-A and endotype-B; proteins in red have a significant adjusted p-value and fold change. **(B)** t-SNE plot representations for endotype-A and endotype-B patients with ISS>15, showing average expression of all proteins for these groups relative to the mean in all patients. **(C)** Density plots of four neuronal guidance proteins (UNC5B, SEMA7A, EPHB6, RGMB) that are differentially expressed between endotype-A and endotype-B individuals (ISS>15). **(D)** PCA of the Shap values generated by the model utilizing the 23 ML-derived proteins to predict critical injury (i.e., ISS>24), with patients suffering various adverse clinical outcomes indicated. Endotype-A and endotype-B patients are shown. **(E)** Table displaying adverse outcomes for each endotype. For each individual outcome, the table shows both the absolute number and percentage, plus fold-change and odds ratio.

## Endotype-B patients have greater likelihood of adverse clinical outcomes

The elevated protein level and striking NGP signature in the hyperacute plasma of endotype-B patients suggest significant physiological perturbations (regardless of their ISS), that may lead to greater probability of adverse clinical outcomes, such as death, prolonged MODs, extended stay in the ICU, or infection. To assess this, we highlighted individuals with these outcomes on PCA plots of Shapley values for the ML_(23)_^ISS>24^ model (**Fig. 4D**). Endotype-B patients had significantly worse clinical outcomes than endotype-A patients, having odds ratios of 34.0 (C.I. 4.5 - 259.3) for early deaths, 9.8 (C.I. 1.1 – 84.6) for late deaths, 15.4 (C.I. 4.4 – 53.2) for prolonged MODS, 7.0 (C.I. 3.9 – 12.6) for ICU stay greater than 7-days, and 3.1 (C.I. 1.8 – 5.3) for infection (**Fig. 4E**). Thus, plasma protein levels, measured within 2hr of major trauma, identify two serious-injury endotypes that do not segregate by ISS. Individuals with endotype-B (compared to endotype-A), have elevated plasma protein levels and a profound neuronal guidance protein signature, and have significantly greater likelihood of adverse clinical outcomes in the days that follow their injury.

## Endotype-B patients with good outcomes display a “protective” hemostatic signature

Although endotype-B individuals have significantly increased likelihood of adverse clinical outcomes, ∼50% of these individuals nonetheless recovered well (**Fig. 5A**). To ascertain whether endotype-B individuals with bad *versus* good clinical outcomes had important physiological differences, we next assessed differential protein levels between these groups. A relatively small number of proteins (∼30) were significantly differentially represented (**Fig. 5B**), a sizable fraction of which were implicated in hemostasis (**Fig. 5C**). This “protective” signature was characterised by higher levels of PLG (plasminogen), SERPINF2 (alpha-2 anti-plasmin), SERPINC1 (antithrombin III) and SERPINA5 (protein C inhibitor), the latter correlating well with lower levels of PROC (activated protein C) (**Fig. 5D and 5E**). By contrast, in Endotype-B patients with adverse clinical outcomes this pattern was reversed. Thus, trauma patients that display a pronounced neuronal guidance protein signature, coupled with a low-coagulation/hyper-fibrinolytic hemostatic profile, in their blood plasma in the immediate aftermath of serious injury, progress down unfavourable clinical trajectories and suffer multiple adverse clinical outcomes.

**Figure 5.**
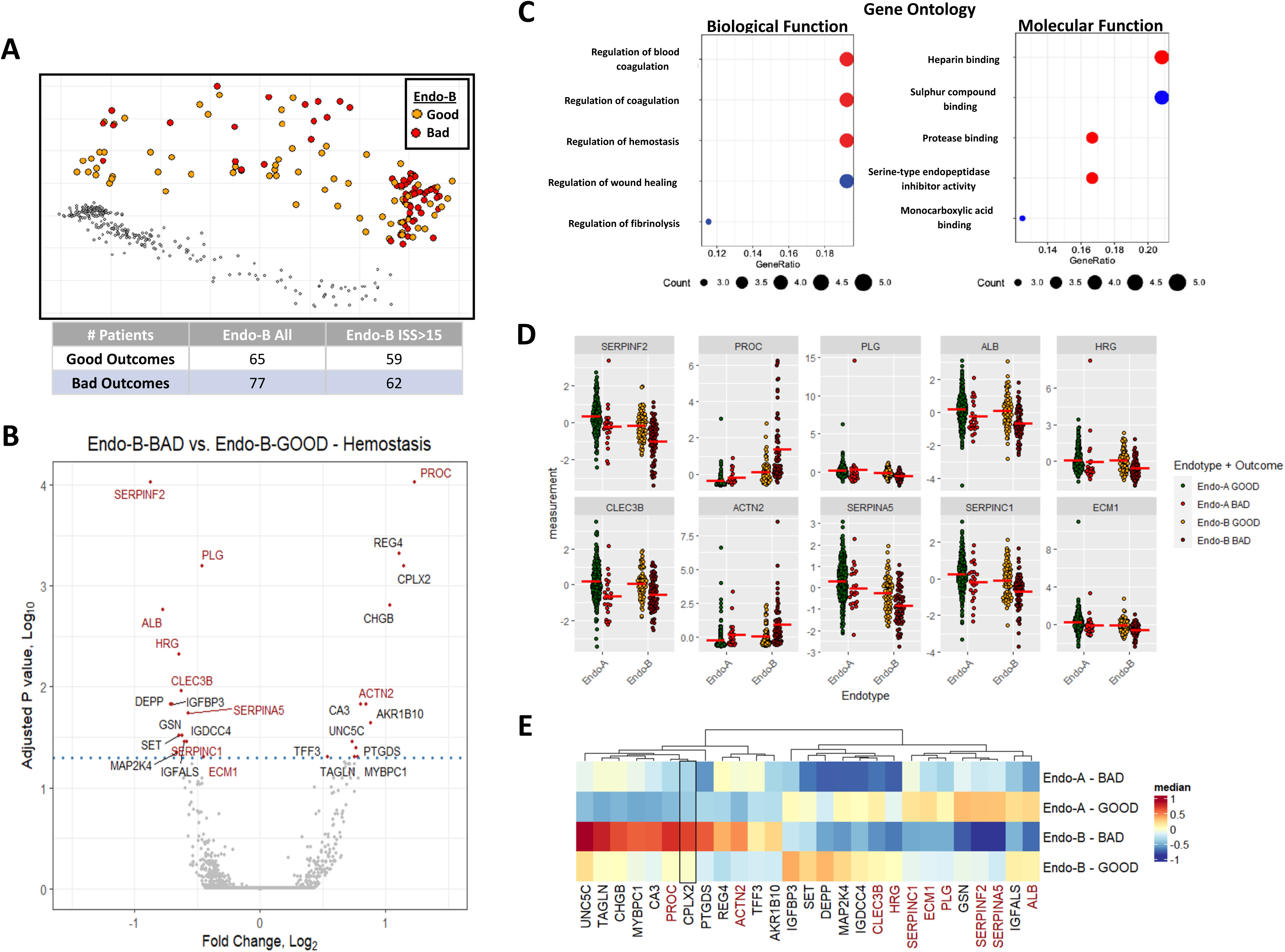
Endotype-B patients with good outcomes display a “protective” hemostatic signature. **(A)** PCA of the Shap values generated by the model utilizing the 23 ML-derived proteins to predict critical injury (i.e., ISS>24). Endotype-B individuals with good (yellow), or bad (red) clinical outcomes are indicated. **(B)** Volcano plot of differentially represented proteins between endotype-B with bad clinical outcomes versus endotype-B patients with good clinical outcomes; proteins above the dashed line have a significant adjusted p-value and fold change. **(C)** Gene ontology analysis for biological and molecular function for proteins identified as significantly different in panel (B). **(D)** Scatter plots showing the level of the 10 hemostatic proteins identified in panel (B) in endotype-A or endotype-B patients with good or bad clinical outcomes. **(E)** Heatmap showing comparative levels for all differentially represented proteins identified in (B) in endotype-A or endotype-B patients with good or bad clinical outcomes. Bad clinical outcomes include death, prolonged MODS (>7 days) or length of stay in the ICU > 7 days.

## Discussion

It has been unclear whether contrasting clinical trajectories of trauma patients with subjectively similar injuries, are a consequence of divergent host responses, or an inability to accurately assess injury severity. Here, we sampled the plasma proteome of 414 trauma patients within 2hr of injury to generate an objective plasma-injury “score”. This contrasts with other studies, in which proteomic analyses are primarily used to predict adverse clinical outcomes (*14, 15*). Our machine-learning approach identified 23 plasma proteins that accurately predict critical injury when benchmarked against ISS, the current gold-standard (*4, 5*). Notably, this identified patients with discordant plasma-injury score/ISS, intimating that these individuals were significantly more or less injured than clinical assessment had suggested. Moreover, we found no evidence for a marked step-change in proteomic signature that would indicate a threshold-breaking transition from a healthy state to one linked to adverse clinical outcomes. Instead, a subset of the 23 proteins correlated directly with ascending ISS, and thus featured prominently in the top differentially represented proteins between critically injured (ISS>24) and low-ISS (ISS<3) individuals. Many appeared to be tissue-specific intracellular proteins (e.g., ADSSL1) that are presumably liberated by extensive tissue destruction. Indeed, these proteins could serve as an objective indicator of injury-induced tissue damage.

Our ML approach also identified two serious-injury endotypes; individuals linked by distinct proteomic phenotypes with common underlying mechanistic features. Both endotypes contained patients from all ISS categories, although endotype-A included the majority of patients with lower ISS. Consistent with this, clinical outcomes for endotype-A patients were generally good, even for those with high ISS, with only 5 individuals (from 271) either dying or developing prolonged organ dysfunction. By contrast, endotype-B patients suffered much worse clinical outcomes, for example being fifteen times more likely to suffer prolonged multiple organ dysfunction syndrome than endotype-A individuals.

A striking feature of endotype-B patients was elevated plasma protein levels. This was also reported in a notable proteomic investigation of 112 severely injured patients enrolled in the Prehospital Air Medical Plasma (PAMPer) trial (*15, 16*). Although this cohort differed from ours, as it targeted individuals with haemorrhagic shock and included 64 patients with traumatic brain injury, it nonetheless suggests that raised plasma protein levels are common in severe trauma. This “systemic storm” was interpreted as a massive release of proteins, by both active and passive mechanisms (*15*). Alternatively, protein levels could also increase due to significant fluid (but not protein) loss from the microvasculature. Trauma-induced endotheliopathy, with loss of barrier integrity (and edema), has been noted as early as ten minutes after serious injury and generally correlates with poor clinical outcomes (*17*).

A defining characteristic of endotype-B patients was elevated representation of neuronal guidance proteins (NGPs) (*18*). These included the netrin (UNC5B, UNC5C, UNC5D and FLRT2), semaphorin (SEMA7A), ephrin (EPHB4 and EPHB6), and RGM (RGMB) systems (*11*). Although historically recognised for driving neuronal patterning during embryonic development, recent studies have also demonstrated profound roles for NGPs in both immune system (*12*), and endothelial biology (*19*). Mechanoreceptor-mediated detection of unfavourable hemodynamic environments (e.g., oscillatory sheer stress in reperfusion injury (*13*)), can up-regulate NGPs (*20, 21*), that have both protective (e.g., Netrin-1 via UNC5B (*22*)), and stimulatory (e.g., SEMA7A via PLXNC1 (*21*)), effects on barrier function and endothelial cell activation. NGPs can also regulate migration of neutrophils and monocytes across endothelial layers (*21, 23*), and mediate thromboinflammatory activation of platelets, (e.g., SEMA7A via GPIb (*24*)). Thus, this evolutionary ancient system of attractive and repulsive cues has been co-opted to modulate many facets of endothelial and immune cell biology that together regulate acute inflammatory and thrombotic events (*12, 13*). Notably, endotype-B individuals have proteomic perturbations REFlecting massive mobilisation of the NGP network, that includes receptors (e.g., UNC5B & EPH6B), normally tethered on surface membranes. This suggests dysregulation and/or dysfunction of the NGP system, driving thromboinflammatory events that could push endotype-B patients down unfavourable clinical trajectories.

Despite sharing an elevated NGP signature, not all endotype-B patients progress along unfavourable clinical paths. Indeed, differential protein analysis revealed a striking anti-coagulant/pro-fibrinolytic signature in endotype-B individuals that subsequently developed clinical complications. This featured high levels of activated protein C (APC) that correlates with trauma-induced coagulopathy (TIC) and poor prognosis (*9, 25*). Protein C is bound by endothelial protein C receptor (EPCR) and is activated by the thrombin/thrombomodulin complex. APC exerts its anti-coagulant function via inactivation of clotting factors V and VIII that are required for thrombin production. Additionally, APC inactivates SERPINE1 (plasminogen activator inhibitor-1), that allows tissue plasminogen activator to generate plasmin to breakdown fibrin clots. Lower levels of plasminogen (PLG), protein C inhibitor (SERPINA5), and alpha-2 antiplasmin (SERPINF2), in endotype-B patients with poor outcomes, are also consistent with this hypo-coagulation/hyper-fibrinolysis state.

Collectively, these data demonstrate the utility of the plasma proteome as an immediate indicator of injury severity. We identify an “at-risk” severe-injury endotype, characterised by a pronounced NGP signature that is a signal-bearer and mediator of profound vasculature stress, endothelial dysfunction, and immune cell activation. Those individuals that react to this vascular challenge by entering an anti-coagulation/pro-fibrinolysis state, likely suffer further vascular collapse, systemic edema, and damaging immunopathology that drives these patients down poor recovery trajectories with multiple clinical complications. Thus, these new insights into the mechanisms underpinning the immediate host response to severe trauma provide tractable new targets for potential therapeutic intervention to improve recovery rates in the critical care setting.

## Data Availability

All data produced in the present study are available upon reasonable request to the authors and will be shared publicly on journal publication

## Acknowledgments

We thank all laboratory members for discussions, especially C. Cabrera for bioinformatic advice.

## Author contributions

K.B, M.R.B. and D.J.P. conceived the project, acquired the funding, and supervised the project. S.M., G.H-M., and J.R. performed the bioinformatic analyses. D.J.P. wrote the manuscript. All authors conceptualised the data, and reviewed and edited the manuscript.

## Data and materials availability

All data are available in the main text or the supplementary materials unless stated here.

## Supplementary Materials

### Materials and Methods

#### Patients

Patients were from the Activation of Coagulation and Inflammation in Trauma (ACIT-II) study, that is a prospective platform cohort study evaluating aspects of coagulation and inflammation in trauma patients (NHS REC: 07/Q0603/29) . Patients at The Royal London Hospital were enrolled into ACIT-II in the emergency department. Admission blood samples (taken within 2h), and physiology data were collected on arrival, and further clinical data were collected daily over the following 28-days or until discharge from hospital or death. The Injury Severity Score (ISS) for each patient was calculated once all injuries had been fully assessed and documented. Patients here (414), were enrolled between Feb-08 and Mar-19. Only patients with blunt trauma were included, and those with penetrating injuries or Traumatic Brain Injuries (TBI) were excluded. Patients were selected to represent a range of injury severities (ISS 0-3 = 73, ISS 4-8 = 77, ISS 9-15 = 79, ISS 16-24 = 78, ISS>24 = 108).

#### Proteomics

For proteomic analysis blood was collected in EDTA tubes (P100 Blood Collection System, BD Biosciences), centrifuged at 2810g at room temperature, aliquoted, and stored at -80C. Samples were later thawed, re-aliquoted, re-frozen and transported for external analysis using the SOMAScan proprietary proteomics platform . SOMAScan uses synthetic aptamers in a highly multiplexed assay to detect ∼5000 proteins in a single sample. SOMAScan proteomics data is provided in Relative Fluorescence Units (RFU).

#### Statistical testing and modelling

The data was standardised by removing the mean and scaling to unit variance. The degree of association between the Injury Severity Score (ISS) and continuous variables in our clinical dataset and proteomic dataset was calculated using Pearson and Spearman coefficients (see table).

#### Differential expression analysis

Relative protein abundance was provided by Somalogic. Data was delivered in microarray format (arbitrary fluorescence units). To estimate statistically-significant differences in plasma protein abundance between groups we performed differential expression analysis using *limma* package in R. Briefly, scaled and centred data were fit to a linear model with empirical Bayes moderation and Benjamini-Hochberg correction was applied to adjust significance for multiple comparisons (adjusted p-value = False Discovery Rate (FDR)).

#### Pathway analysis

Characterisation of biological processes and molecular functions was performed by over-representation pathway analysis. Statistical significance (enrichment) was calculated by hypergeometric test against a specific set of pathway databases (Gene Ontology, KEGG, Reactome and WikiPathways) in R (using DOSE, clusterProfiler, ReactomePA packages) or in Python/R using enrichR. In all cases, somamers were matched to their gene EntrezID equivalent with duplicates and NAs removed. The probable gene universe for pathway discovery was trimmed to fit the single genes present in the SomaLogic array.

#### Protein clustering

Clustering was performed using the KMeans function from Sklearn (Python) to identify proteins belonging to common pathways. We found that 20 was the ideal number of clusters for our dataset. To show protein expression changes across different severity groups we calculated the difference between the mean expression of proteins across all patients and the mean expression of a particular group. Finally, to visualise and characterise pathway variation across different groups, we overlaid this data on a t-SNE plot, allowing a non-linear low dimensional representation of the data. We repeated this process for all groups of interest. To accompany the t-SNE plot, we created a table summarising the most important pathways associated with each cluster. To extract the pathways from Enrichment databases (EnrichR) we used the Python package enrich_omics.

#### Machine Learning Framework

To evaluate protein importance, we developed a framework composed of ensemble classifiers training and testing at thresholds in the range of ISS>24 to ISS>33. To implement the individual models of our framework, we used the Python library XGBoost with a learning rate of 0.2 and adjusted weights for class imbalance. We trained the model for 35 epochs. Model hyperparameters were estimated using a Grid Search and train/test/validate splits for both the protein search step and the individual model. We tested learning rates in a range 0.1-0.5 and tree depths in a range 1-4. After every model iteration, we extracted the most important features (using F-score as metric) which were then merged to create a “consensus set” of 23 proteins. These features were subsequently used to train a model classifying patients with ISS higher or lower than 24. They were subsequently explained using SHAP’s “Summary Plots” and “Force Plots” allowing us to obtain global (model-level) and local (or patient-level) explanations of our models . The importance of proteins in the two final models and their effect on the prediction were shown as mean SHAP values on a summary plot and as individual SHAP values on Force Plots.

---

**Supplementary figure 1.**
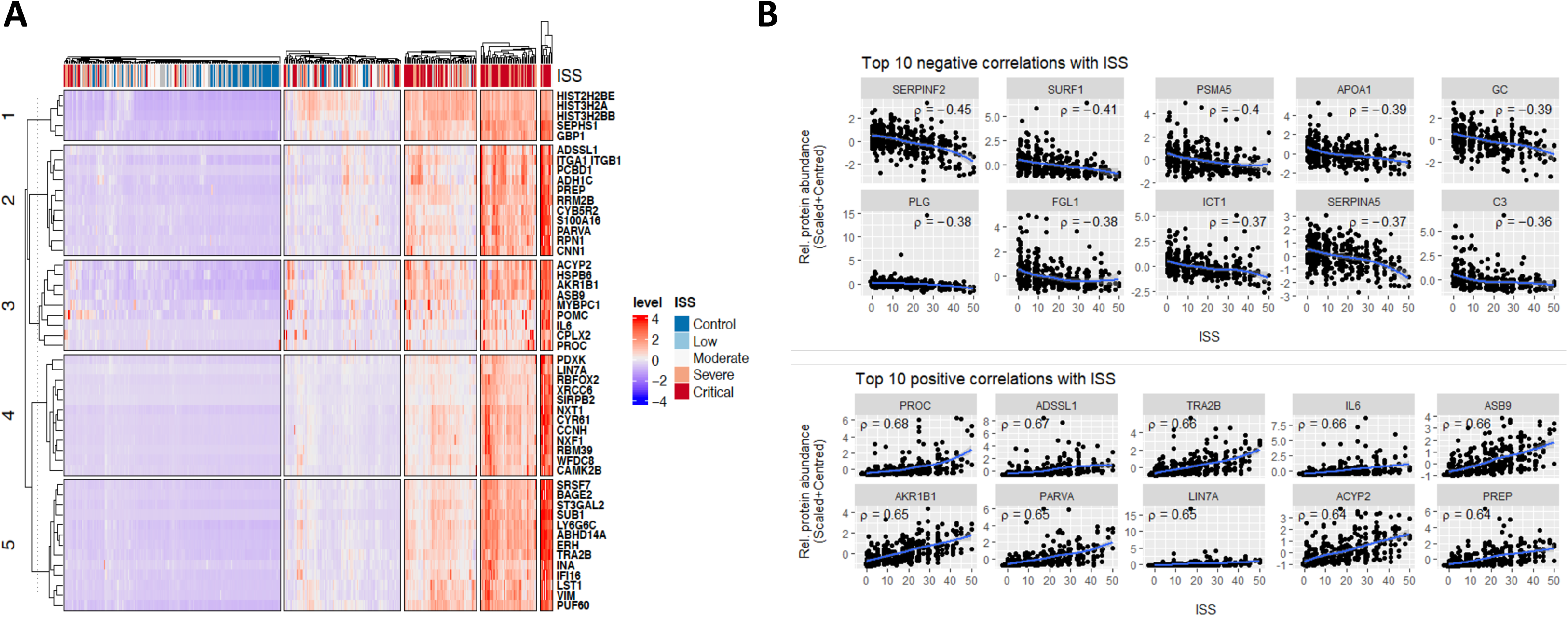
(**A**) Heat map showing the top 50 proteins whose levels correlate with injury severity score (ISS). (**B**) Scatter plots for the top 10 proteins with negative (top) or positive (bottom) correlations with ISS.

**Supplementary figure 2.**
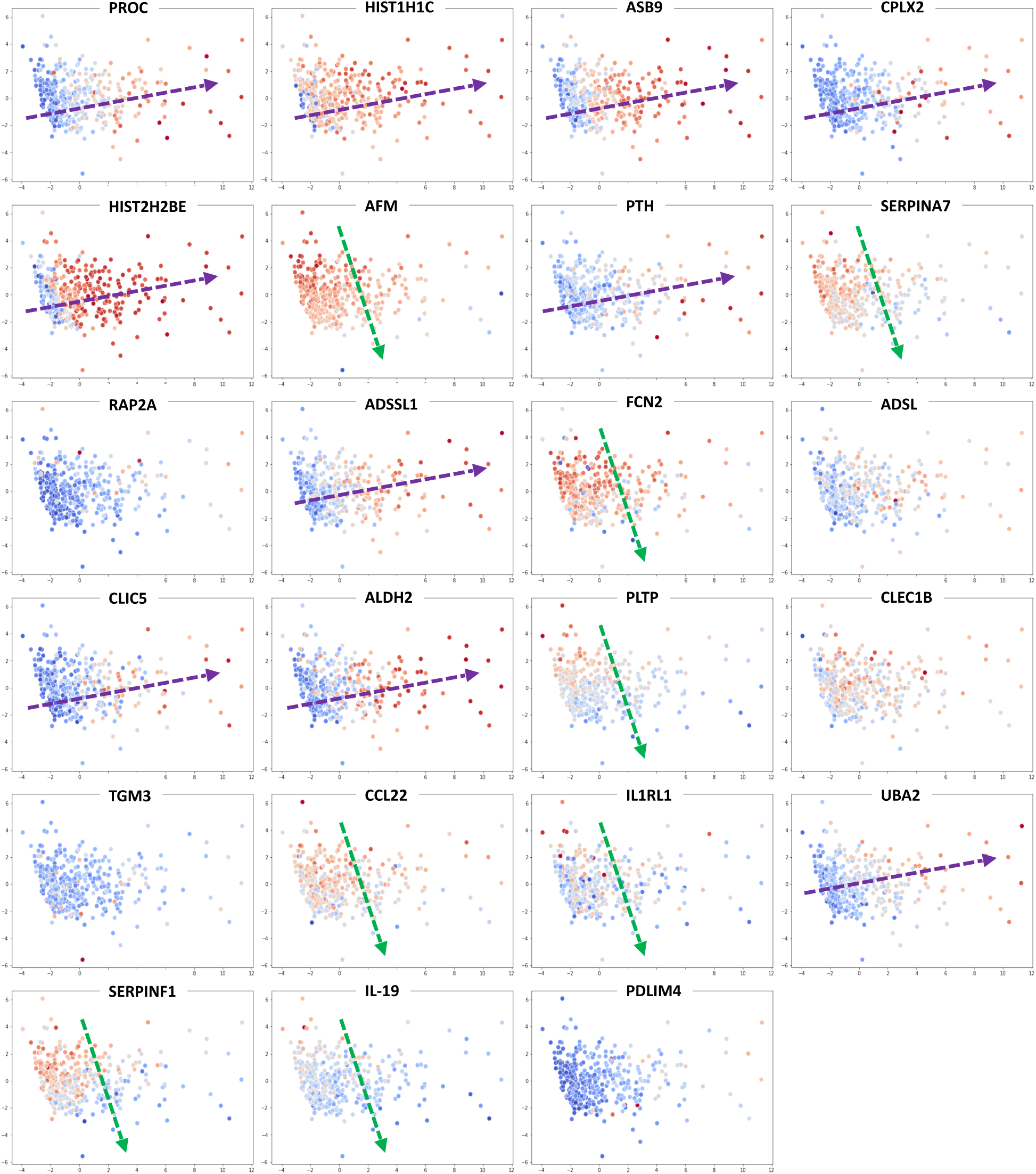
PCA based on expression of the 23 M-L-derived proteins in our patient cohort, coloured by relative expression of each protein. Red indicates higher protein expression. Arrows shows “direction” of increasing or decreasing protein expression.

**Supplementary figure 3.**
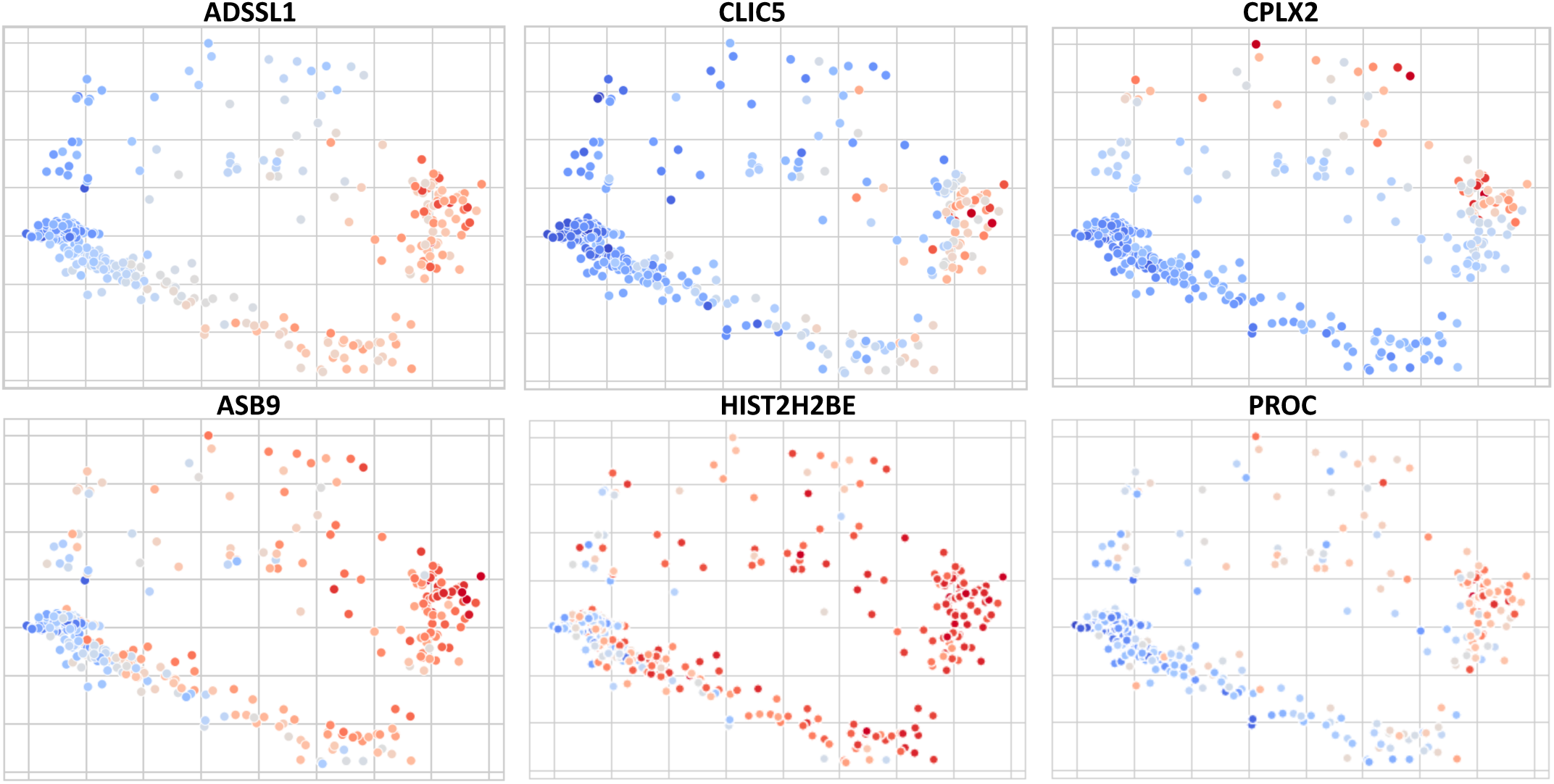
PCA of the Shapley values generated by the model utilizing the 23 M-L-derived proteins to predict critical injury (i.e., ISS>24), showing expression of the six most dominant proteins. Red denotes higher expression.

**Supplementary figure 4.**
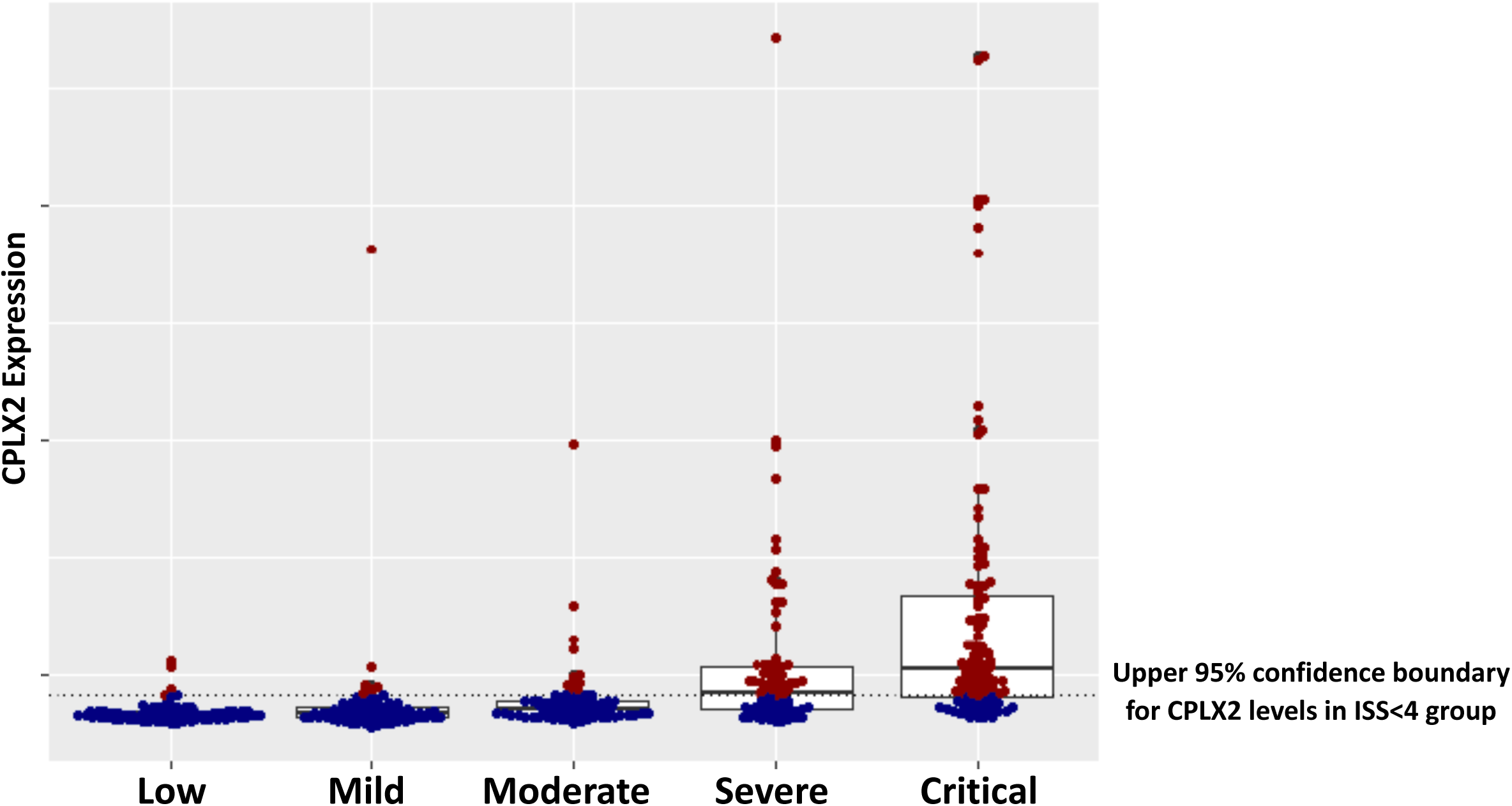
Scatter plot showing endotype-A (blue) and endotype-B (red) patients for each ISS group.

**Supplementary figure 5.**
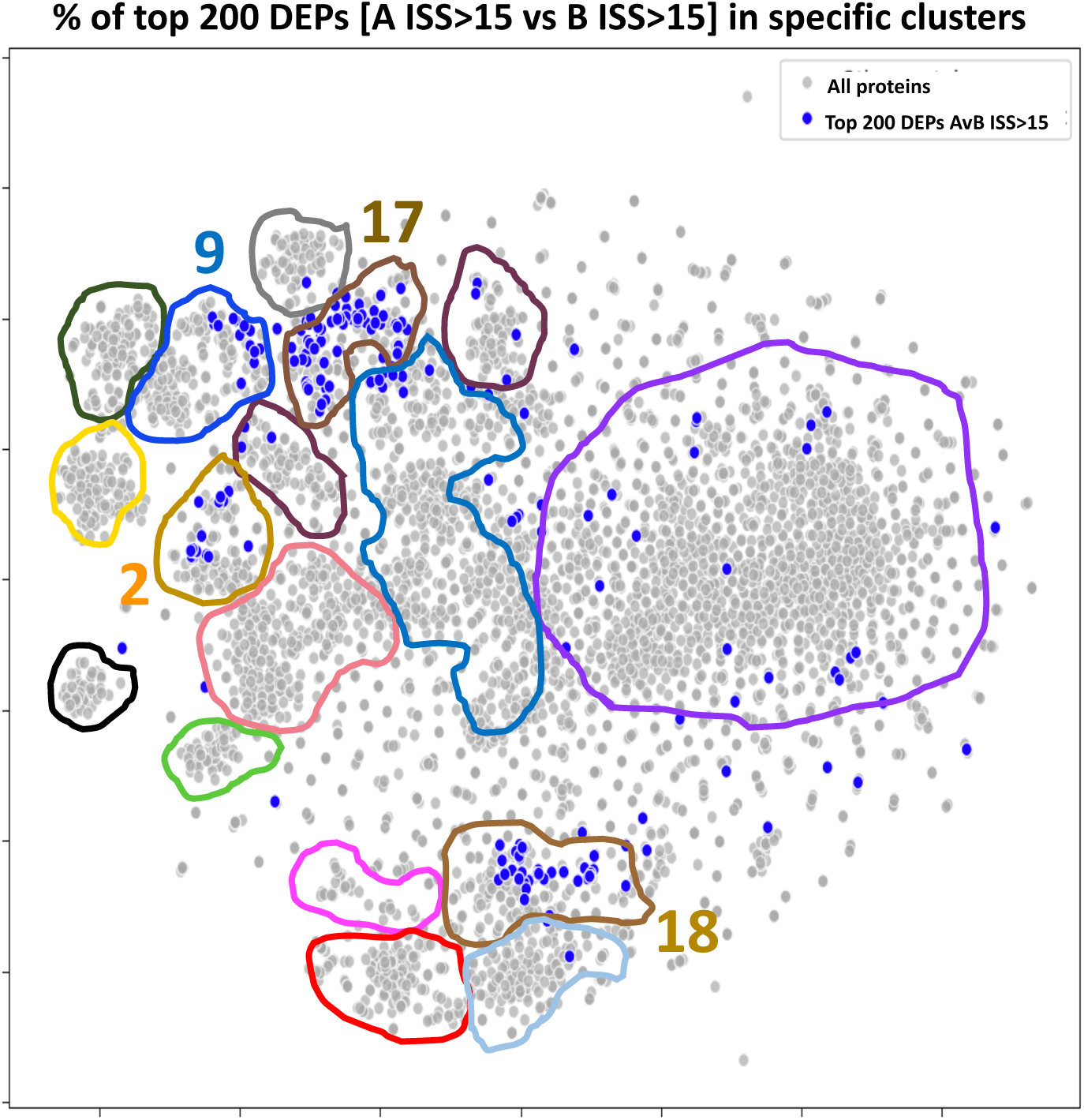
Top 200 differentially expressed proteins from comparison of patients with ISS>15 from endotype-A and endotype-B, overlaid onto a t-SNE plot showing proteins clustered (K-means) based on protein expression in our 414-patient cohort.

**Supplementary figure 6.**
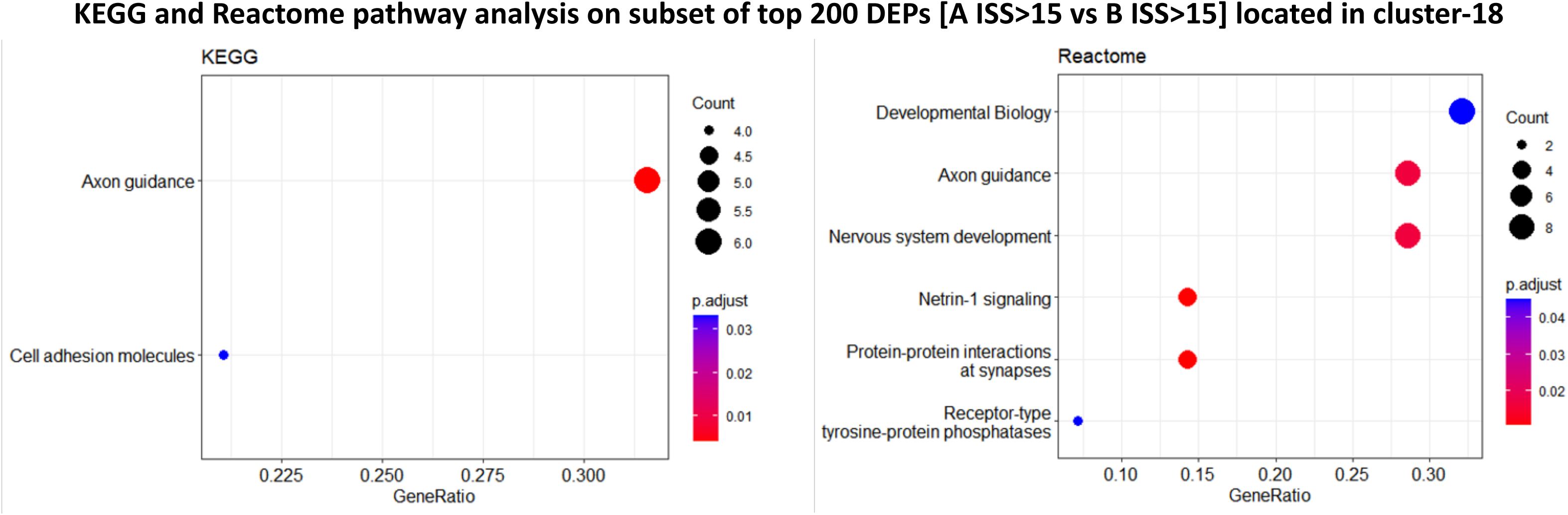
KEGG and Reactome pathway analysis on top 200 differentially expressed proteins between patients with ISS>15 from endotype-A and endotype-B located in the boundaries of cluster 18 from the t-SNE plot shown in Figure 3. (

**Supplementary figure 7.**
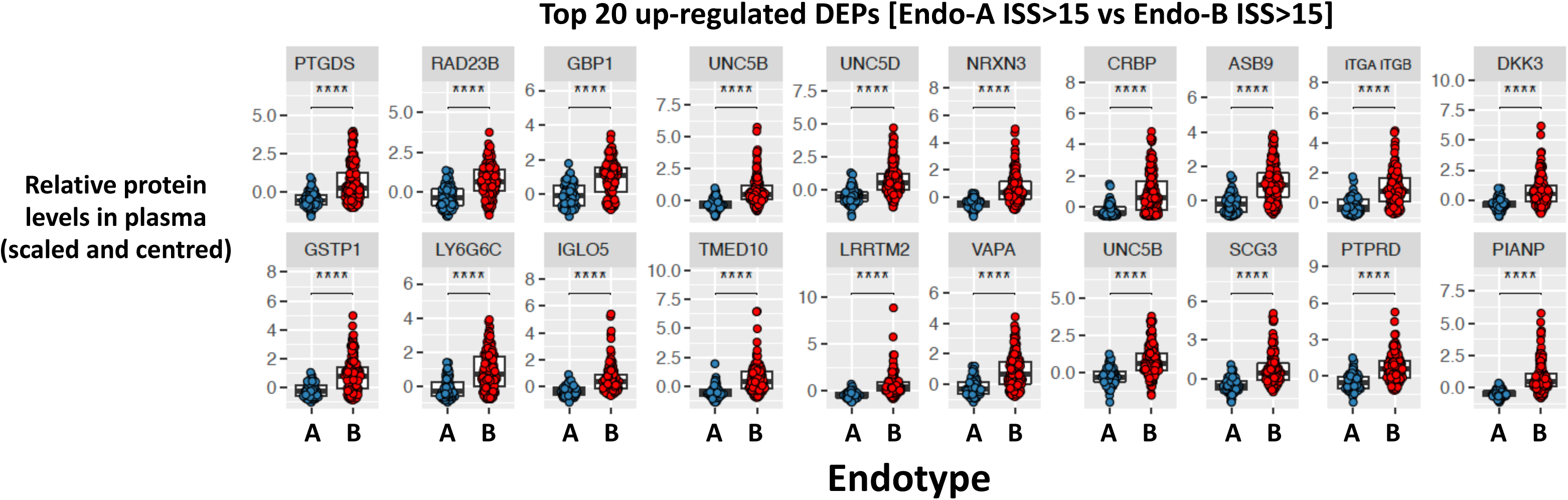
Scatter plot of top 20 up-regulated proteins from (A) showing relative protein levels of scaled and centred dataset.

**Supplementary figure 8.**
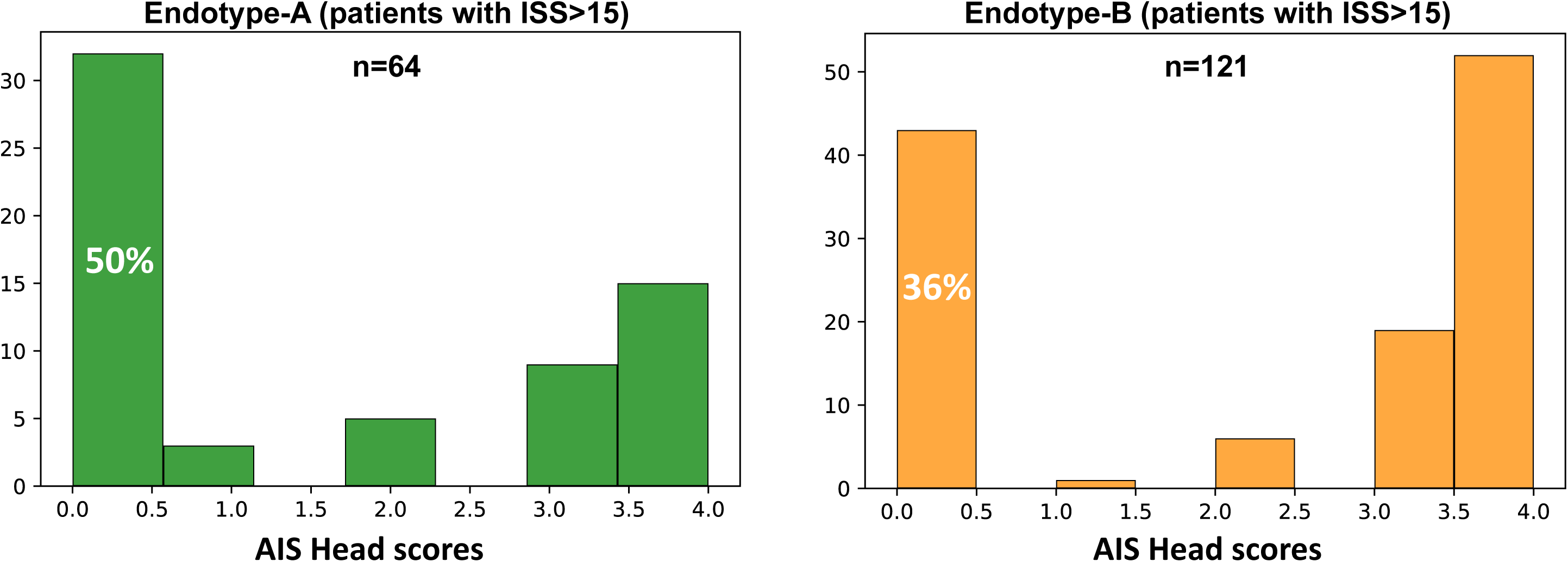
Bar plots showing AIS Head scores for endotype-A patients with ISS>15 (green) and endotype-B patients with ISS>15 (orange). Number of patients shown in each plot is indicated.

**Supplementary table 1.** Cohort data. Table displaying clinical data at admission for patients separated into ISS categories; low (ISS 0-3), mild (ISS 4-8), moderate (ISS 9-15), severe (ISS 16-24), and critical (ISS 25-75).

|  | Iss<br>range | Patients<br>count | Injury to<br>baseline<br>time | Systolic<br>Blood<br>pressure<br>(<120mmHg) | Baseline<br>wcc<br>(4.5-11<br>10<sup>9</sup>/L) | Baseline<br>lactate<br>(0.5-2.2mmol/L) | Baseline<br>base<br>deficit(-2/+2<br>mEq/L) | Length<br>of stay | Length<br>of stay<br>>14d | Mortality<br>rate | Infection<br>rate | Short<br>Mods | Prolonged<br>Mods | Any<br>Mods | No<br>Mods |
| --- | --- | --- | --- | --- | --- | --- | --- | --- | --- | --- | --- | --- | --- | --- | --- |
| **Low** | 0-3 | 73 | 86.0 ±<br>20.3 | 137.6 ± 25.9 | 10.0 ±<br>4.0 | 2.3 ± 2.4 | -0.1 ± 3.3 | 1.7 ±<br>2.3 | 0 | 0/73 (0) | 1/65<br>(0.02) | 0 | 2 | 2 | 71 |
| **Medium** | 4-8 | 77 | 84.4 ±<br>20.4 | 142.2 ± 25.1 | 10.8 ±<br>4.1 | 2.1 ± 1.8 | 0.4 ± 2.8 | 7.0 ±<br>9.4 | 11 | 0/77 (0) | 5/71<br>(0.07) | 1 | 4 | 5 | 72 |
| **Moderate** | 9-15 | 79 | 87.3 ±<br>22.1 | 133.9 ± 24.5 | 12.7 ±<br>5.7 | 2.6 ± 2.3 | 0.5 ± 3.1 | 15.5 ±<br>16.8 | 30 | 2/79<br>(0.03) | 14/71<br>(0.2) | 3 | 8 | 11 | 68 |
| **Severe** | 16-24 | 77 | 88.4 ±<br>26.5 | 134.5 ± 29.4 | 16.2 ±<br>6.2 | 2.5 ± 2.4 | 1.8 ± 3.6 | 18.2 ±<br>15.7 | 36 | 5/78<br>(0.06) | 14/61<br>(0.23) | 4 | 14 | 18 | 59 |
| **Critical** | 25-50 | 108 | 96.5 ±<br>22.8 | 124.4 ± 33.6 | 17.1 ±<br>7.2 | 3.1 ± 2.6 | 3.4 ± 6.3 | 26.7 ±<br>27.4 | 66 | 16/108<br>(0.15) | 32/73<br>(0.44) | 16 | 35 | 51 | 57 |

**Supplementary table 2.** Table showing the most important features (proteins) identified by our model at different ISS thresholds. Importance was calculated using the F-score.

|  | Prot #1 | Prot #2 | Prot #3 | Prot #4 | Prot #5 | Prot #6 | Prot #7 |
| --- | --- | --- | --- | --- | --- | --- | --- |
| Threshold at 24 | PROC | HIST1H1C | N/A | N/A | N/A | N/A | N/A |
| Threshold at 25 | ASB9 | CPLX2 | HIST2H2BE | AFM | PTH | SERPIINA7 | N/A |
| Threshold at 26 | ASB9 | CPLX2 | AFM | PTH | RAP2A | N/A | N/A |
| Threshold at 27-28 | ADSSL1 | ASB9 | CPLX2 | PTH | FCN2 | RAP2A | ADSL |
| Threshold at 29 | ADSSL1 | CLIC5 | ALDH2 | PLTP | CLEC1B | TGM3 | N/A |
| Threshold at 30-32 | ADSSL1 | CLIC5 | CCL22 | PLTP | IL1RL1 | N/A | N/A |
| Threshold at 33 | ADSSL1 | UBA2 | SERPINF1 | AFM | IL19 | PDLIM4 | CLEC1B |

**Supplementary table 3.** Performance of the 23-protein ML model on training and test datasets. Precision, recall, F1 score and AUC are illustrated as indicators of model performance.

| Score | Train | Test |
| --- | --- | --- |
| AUC | 0.98 | 0.88 |
| Precision (w. avg) | 0.92 | 0.82 |
| Recall (w. avg) | 0.91 | 0.80 |
| F1 score | 0.91 | 0.81 |

**Supplementary table 4.** Table comparing admission data of patients with an actual ISS>24 (bottom row), with patients predicted as having an ISS>24 by the ML_(23)_^ISS>24^ model (top row), from a test cohort of trauma patients.

|  | Counts | Age | Systolic Blood pressure (<120mmHg) | Baseline wcc (4.5-11 10<sup>9</sup>/L) | Baseline lactate (0.5-2.2mmol/L) | baseline base deficit (-2/+2 mEq/L) | length of stay | Mortality | Infection | Short MODS | Prolonged MODS | Any MODS |
| --- | --- | --- | --- | --- | --- | --- | --- | --- | --- | --- | --- | --- |
| Predicted critical | 40 | 42.2 ± 18.2 | 90.1 ± 22.7 | 17.4 ± 7.1 | 3.5 ± 2.5 | 4.1 ± 5.8 | 25.5 ± 20.4 | 6/40(0.15) | 11/25 (0.44) | 7/40 | 10/40 | 17/40 |
| ISS critical >=25 | 33 | 46.4 ± 19.0 | 88.3 ± 21.3 | 15.7 ± 6.6 | 3.1 ± 2.5 | 3.5 ± 5.5 | 29.3 ± 28.4 | 6/33 (0.18) | 8/22 (0.36) | 7/31 | 8/31 | 15/31 |

**Supplementary table 5.** Table showing details of all clusters present on t-SNE plots based on protein expression in our 414 patients. Table displays cluster number, the major pathway and PC associated to that cluster, and whether one of the 23 M-L-derived proteins is present in that cluster.

| Cluster | General description of statistical significant pathways | # Somamers | PC-dom | ML somamers |
| --- | --- | --- | --- | --- |
| Cluster 0 | Syalyltransferase | 4 |  |  |
| Cluster 1 | Platelets | 114 | **both** | **CLEC1B** |
| Cluster 2 | Metabolism | 142 | **PC1** | **ADSSL1, ALDH2** |
| Cluster 3 | Cytokine signalling | 1545 |  |  |
| Cluster 4 | Digestion | 277 |  | CPLX2, TGM3, PDLIM4 |
| Cluster 5 | Neurotransmission | 299 |  |  |
| Cluster 6 | Neuro and immune signalling | 382 |  |  |
| Cluster 7 | Granules | 175 | **PC1** | **HIST1HC** |
| Cluster 8 | Extracellular matrix | 172 | **PC2** | **PLTP, IL19, IL1R1** |
| Cluster 9 | Protein translation | 176 | **PC1** | **UBA2** |
| Cluster 10 | Lipid mediators metabolism | 86 | **PC2** | **CCL22** |
| Cluster 11 | - | 300 |  | PTH, RAP2A, ADSL |
| Cluster 12 | Nucleotide metabolism | 111 |  |  |
| Cluster 13 | Ubiquitination | 61 |  |  |
| Cluster 14 | RNA processing | 524 |  |  |
| Cluster 15 | Cytoskeleton organisation and cell migration (+ TLR and VEGF signalling) | 93 |  |  |
| Cluster 16 | Hemostasis, platelet activation and chemokine signalling | 57 |  |  |
| Cluster 17 | Cellular responses to stress (DNA/RNA/Cytoskeleton) and Estrogen signalling | 189 | **PC1** | **PROC, CLIC5, ASB9, HIST2H2BE** |
| Cluster 18 | Extracellular matrix | 139 |  |  |
| Cluster 19 | Complement and Coagulation | 133 | **PC2** | **AFM, SERPINF1, SERPINA7, FCN2** |

**Supplementary table 6.** Table comparing admission data of endotype-B patients with good versus adverse clinical outcomes.

|  | Count | Sex<br>M/F | Age | ISS | Systolic Blood pressure<br>(<120mmHg) | Baseline wcc (4.5-11<br>10*9/L) | Baseline lactate (0.5-2.2mmol/L) | Baseline base deficit (-2/+2<br>mEq/L) |
| --- | --- | --- | --- | --- | --- | --- | --- | --- |
| Endotype A<br>(ISS>15) | 64 | 97/24 | 44.0 ±<br>15.2 | 25.7 ±<br>8.5 | 132.9 ± 27.8 | 16.0 ± 5.9 | 2.5 ± 2.3 | 1.5 ± 3.4 |
| Endotype B<br>(ISS>15) | 121 | 53/11 | 44.6 ±<br>19.7 | 29.0 ±<br>9.0 | 126.6 ± 34.1 | 17.1 ± 7.2 | 3.1 ± 2.6 | 3.4 ± 6.0 |

